# Modelling the first wave of the COVID-19 epidemic in the Czech Republic and the role of government interventions

**DOI:** 10.1101/2020.09.10.20192070

**Authors:** Ondřej Májek, Ondřej Ngo, Jiří Jarkovský, Martin Komenda, Jarmila Rážová, Ladislav Dušek, Tomáš Pavlík

## Abstract

In the Czech Republic, the first COVID-19 cases were confirmed on 1 March 2020; early population interventions were adopted in the following weeks. A simple epidemiological model was developed to help decision-makers understand the course of the epidemic and perform short-term predictions. In this paper, we present the use of the model and estimated changes in the reproduction number (decrease from > 2.00 to < 1.00 over March and April) following adopted interventions.

## INTRODUCTION

More than 27 million of COVID-19 cases and over 800 thousand deaths have been reported globally so far.[1] Population interventions including restrictions limiting public gatherings and social contact have proved crucial in the fight against COVID-19.[2] In the Czech Republic, first COVID-19 cases were confirmed on 1 March 2020. A series of early measures was adopted over the following weeks in accordance with the Public Health Protection Act and the Act on the Security of the Czech Republic (Table 1), leading to rather favourable results after the first wave of epidemic (87 cases and 3 deaths per 100,000 population at the end of May, compared to 269 cases and 32 deaths per 100,000 population in the entire EU/EEA and UK).[3]

**Table 1.** Overview of interventions adopted by the Czech Government against COVID-19

| Date | Confirmed COVID-19 cases | Confirmed COVID-19 cases per 100,000 | Restrictive measure implemented |
| --- | --- | --- | --- |
| 9 March 2020 | 38 | 0.36 | Ban on hospital and retirement home visits |
| 11 March 2020 | 94 | 0.88 | Primary, secondary, and tertiary school closure |
| 12 March 2020 | 116 | 1.09 | The nationwide state of emergency declared |
| 14 March 2020 | 189 | 1.77 | Ban on entry into the Czech Republic for all foreigners coming from (coronavirus) high-risk countries<br>Ban on retail sales and the sales of services in business premises |
| 15 March 2020 | 298 | 2.80 | Ban on entry into the Czech Republic for all foreigners |
| 16 March 2020 | 383 | 3.60 | Ban on the free movement of people at the national level with exceptions including travelling to work, running businesses, visiting doctors, necessary visits to family members, shopping for grocery, drugs, and fuel |
| 19 March 2020 | 765 | 7.18 | Mandatory use of respiratory protective equipment in the public |

A simple epidemiological model was developed at the Institute of Health Information and Statistics of the Czech Republic to help decision-makers understand the course of the epidemics including an estimation of the effective reproduction number,[4] and to facilitate short-term predictions. In this paper, we present results of COVID-19 epidemic modelling in a series of six model editions used by decision makers and published in weekly intervals during the first wave of epidemic in late March and April, and estimate the change of the effective reproduction number following the implementation of restrictive measures.

## DESCRIPTION OF THE EPIDEMIOLOGICAL MODEL USED

In the Czech Republic, only selected laboratories are allowed to perform testing. Test results are then, with minimum delay, reported to the central Information System of Infectious Diseases (ISID), and subsequently validated by the respective regional public health authority. Therefore, ISID allows us to quickly obtain and analyse key data for evaluation of the course of epidemics and to publicly share the current status.[5] Data on basic epidemiological characteristics (cumulative confirmed cases, active cases, incidence, etc.) are available as open data (on-line at https://onemocneni-aktualne.mzcr.cz/api/v2/covid-19).

We developed an original epidemiological model, maintaining the simplicity of statistical models while also considering the mechanics of transmission, which allowed us to better understand the course of the epidemics and to produce more realistic predictions.[6] Our model uses classical S(E)IR approach[7, 8] with the following compartments: S (susceptible), I (infected, set of compartments), R_subcl_ (subclinical cases) and R (removed, laboratory-confirmed COVID-19 cases; see Figure 1).

**Figure 1:**
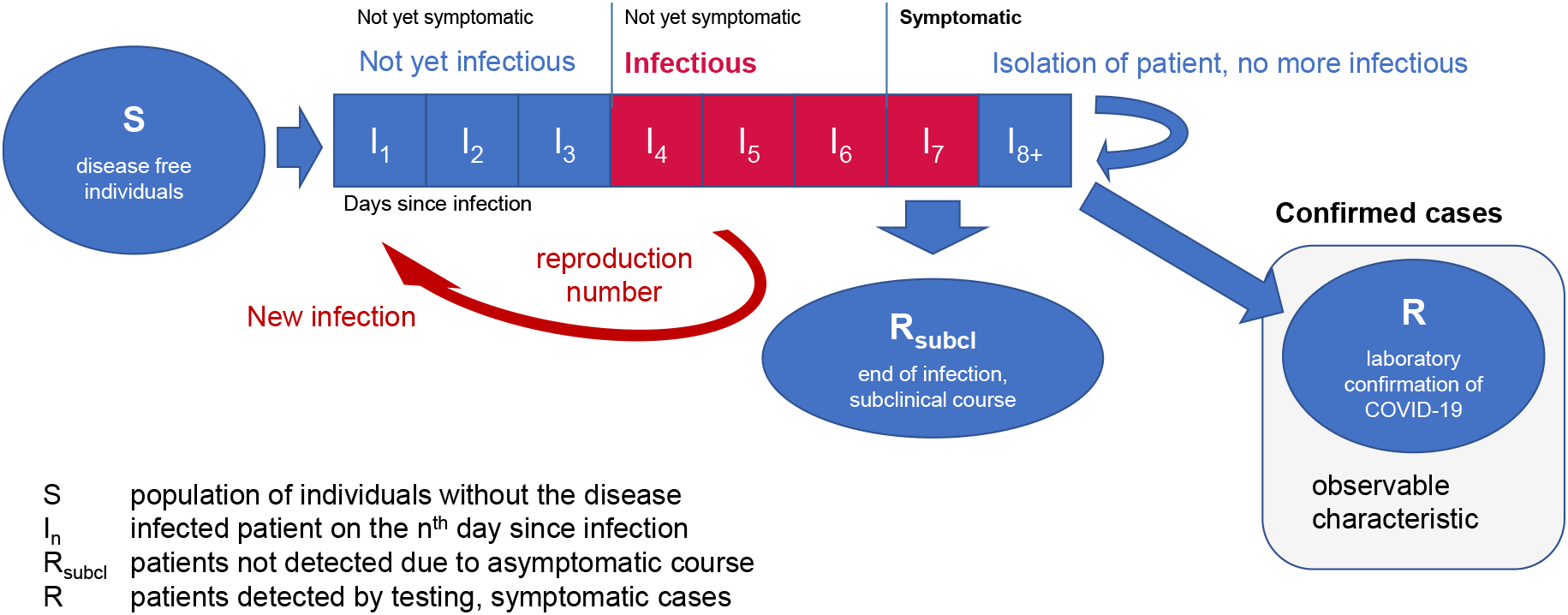
Schema of the epidemiological model

Individuals identified as cases imported from abroad as well as individuals infected within the community (in line with the estimated reproduction number) enter the state I_1_. They stay in the individual compartments I_1_ to I_7_ always for one day. Individuals in states I_4_ to I_7_ can infect others, the number of newly infected individuals depends on the reproduction number. To be able to consider the testing effectiveness (i.e. the delay between referring the patient for testing and the availability of test results), the average length of stay in the I_8+_ compartment was calibrated to the ISID data. For simplicity, it was assumed that the isolation or end of patients’ infectiousness always comes on the second day after the onset of symptoms, limiting further patient’s infectiousness. In line with the general testing policy applicable in the Czech Republic in the respective time period, testing was assumed only in symptomatic individuals. It was assumed that 10 % of infectious individuals were subclinical and would not be included in the ISID statistics (see Table 2 for the complete set of parameters).

**Table 2:** Variables and parameters used in the epidemiological model.

| Variable/parameter | Value | Source | Use in model |
| --- | --- | --- | --- |
| Number of confirmed COVID-19 cases | dataset from ISID | ISID | target value for model calibration |
| Incubation period | 6 days | assumption based on the literature, e.g.[9] | considered in model structure |
| Period of infectiousness within the incubation period | 3 days | assumption based on the serial interval considerations, e.g.[10] | considered in model structure |
| Proportion of subclinical cases | 10 % | assumption based on the literature, e.g.[11] | model parameter, assumed |
| Testing effectiveness: the mean time from onset of symptoms to testing | 7.63 days up to 22/3<br>2.49 days since 31/3<sup>1</sup> | ISID | model parameter, calibrated |
| Number of cases initially imported from abroad | dataset from ISID, 28/3, extrapolated | ISID | setting of the initial epidemic dynamic |

ISID Information System of Infectious Diseases
<sup>1</sup> actual parameter values from the model edition from 30/4; for the period between 22/3 and 31/3, the parameter value was linearly interpolated

Values of selected parameters were calibrated to available ISID data, namely to the cumulative number of confirmed COVID-19 cases. Sum of squares was the metric used for the description of the goodness of fit; except for the first edition, where the manual parameter search was used, the parameter values were derived using the random search method; details are given in the Supplement S1. 5-10% of the best fitting simulations were utilised for parameter estimation (later model editions used 5%). Of these simulations, subset of simulations predicting the highest numbers of cases (see Table 3 for details on the size of these subsets) were used in the early predictions as a precautionary approach. Parameter values were estimated as means of parameter values from those accepted simulations; the reproduction number was also estimated with standard deviations to allow estimation of an indicative 95% confidence interval. The 95% confidence interval bands were subsequently applied as reproduction number values on the recent (retrospective) and on the near future (prospective) period for estimating of a ‘sensitivity interval’, i.e., the predicted interval of the potential numbers of cases at individual target dates (Table 3). Calculations were performed in Microsoft Excel.

**Table 3:** Results of epidemiological modelling, 6 model editions published in weekly intervals (predicted cumulative number of COVID-19 cases)

| Date of publishing | Included data | Prediction: end March | Prediction: end April | Prediction: end May | Evolution of the reproduction number |
| --- | --- | --- | --- | --- | --- |
| 24 March 2020 | By 21 March (1,047 cases) | 3,035 | 15,058 |  | 2.64 (by 11 March)<br>1.84 (12-15 March)<br>1.20 (since 16 March, assumption) |
| 1 April | By 27 March (2,395 cases) |  | 13,965<br>sensitivity interval<sup>1</sup><br>7,201 – 29,880 |  | 2.64 (by 6 March)<br>1.84 (7-11 March)<br>1.32 (12-15 March)<br>1.10, 95% CI 0.87-1.33<sup>1</sup><br>(since 16 March, early estimate) |
| 8 April | By 6 April (4,822 cases) |  | 10,582<br>sensitivity interval<sup>2</sup><br>8,979 – 12,565 |  | ...<br>1.28 (12-15 March)<br>1.02, 95% CI 0.97-1.08<sup>2</sup><br>(since 16 March) |
| 15 April | By 11 April (5,902 cases) |  | 9,949<br>sensitivity interval<br>8,700 – 11,452 |  | ...<br>1.00, 95% CI 0.95-1.05<br>(since 16 March) |
| 23 April | By 21 April (7,033 cases) |  | 7,759<br>sensitivity interval<br>7,370 – 8,250 |  | ...<br>1.00, 95% CI 0.95-1.04 (16-31 March)<br>0.74, 95% CI 0.53-0.96 (since 1 April) |
| 30 April | By 28 April (7,504 cases) |  |  | 8,484<br>sensitivity interval<br>7,923 – 9,677 | ...<br>1.00, 95% CI 0.94-1.07 (16-31 March)<br>0.70, 95% CI 0.49-0.92 (since 1 April) |
| **Observed number** |  | **3,308** | **7,682** | **9,268** |  |

<sup>1</sup> precautionary approach: high-risk decile of best-fitting simulations, confidence interval bands used for the sensitivity interval
<sup>2</sup> precautionary approach: high-risk quartile of best-fitting simulations, confidence interval bands used for the sensitivity interval

## APPLICATION OF THE MODEL

The results of the first model edition were published on 24 March 2020 and predicted a sharp increase till the end of March (more than 3 thousand of cases, compared to 1,047 cases observed on 21 March, Table 3). The real (observed) number of cases indeed exceeded 3 thousand on 30 March.

The basic reproduction number for Czech population was estimated to be 2.64, with a partial decline since 12 March (1.84), reflecting the introduction of the state of emergency and the first nationwide restrictive measures (Table 1). The reproduction number following the broader restrictive measures since 16 March was assumed to be 1.2. In reality, the effect of the restrictive measures was even more dramatic, which lead to a substantial overestimation of the number of cases predicted for the end of April.

Further estimates of the reproduction number lead to downward corrections (1.00 since 16 March, following the restriction of free movement, Table 1). The model edition from 23 April allowed us to estimate a further decrease of the reproduction number since early April (0.74, 95% confidence interval 0.53-0.96). This trend of the declining reproduction number following implemented restrictions is similar to recently published results from Italy.[12]

These data allowed an improved prediction of 7,759 cases at the end of April (sensitivity interval 7,370 – 8,250), which eventually lead to only a slight overestimation of the real observed figure (7,682 confirmed COVID-19 cases). The model published at the end of April predicted 8,484 COVID-19 cases at the end of May, suggesting a controlled course of the epidemic following interventions enacted in March by the Czech government (Table 3). The model and its assumed parameter values underestimated the values observed at the end of May (Table 3, Supplementary Figure S2), which implies a minor increase of the reproductive number in May, following the alleviation of interventions in late April and May.

The basic reproduction number of COVID-19 was previously estimated to be 2.2-3.6.[13, 14] Our simple mechanistic model was able to estimate a similar value. When interpreting the results of the model, we need to bear in mind the substantial parameter and structural uncertainty. In particular, published estimates of the periods of incubation and infectiousness differ among studies (e.g.[9, 15, 16]; we, therefore, needed to assume their values (Table 2) and apply them in the model structure. The values we assumed were consistent with previously published estimates of the serial interval of 4-5 days (e.g.[10]). Besides, substantial uncertainty exists around the proportion of subclinical cases. Nevertheless, the use of an alternative proportion of subclinical cases (i.e., 30% as suggested by Nishihura[17] instead of 10% used originally in our model) did not lead to a substantial change of the prediction during the period of controlled epidemic (7831 vs. 7759 predicted for the end of April using the model published on 23 April). Any new restrictions/alleviations following the prediction represent another important source of uncertainty, as well as the epidemic course under different climatic conditions. Indeed, the model has shown a satisfactory predictive validity; however, great uncertainty is associated with the future values of the reproduction number, which is to a great degree affected by adopted policies and the compliance of the target population.

## CONCLUSION

The described model allowed us to analyse the course of the epidemic, including the estimation of the basic reproduction number, and to perform useful short-term predictions, which facilitated the estimation of the necessary readiness of the healthcare system in the days and weeks after the prediction. The comparison of the predicted and observed numbers of cases is incorporated in the early warning system, which is currently used by policy-makers both on the national and regional levels. The Czech data on COVID-19 epidemic have also demonstrated the potential of early implementation of government measures in slowing the spread of the COVID-19 epidemic.

## Data Availability

Up-to-date Czech COVID-19 epidemiology data are available in a public, open access repository

https://onemocneni-aktualne.mzcr.cz/api/v2/covid-19

## Acknowledgement

We would like to acknowledge all the employees of the Public Health Offices, Institute of Health Information and Statistics, National Public Health Institute and Ministry of Health of the Czech Republic involved in the COVID-19 information support for their great work, which was also vital for preparation of this article. The authors would also like to thank Dr Jaroslav Janošek for his insightful comments.

## Authors’ contribution

Design of the statistical model: OM, ON, JJ, LD, TP; overall concept of the study: LD, OM, TP, JR; data preparation and validation: JJ, MK, JR, LD; calculations: OM, TP, ON, JJ; writing: OM, TP, MK, LD.

## References

1. Center for Systems Science and Engineering (CSSE) at Johns Hopkins University (JHU). COVID-19 Dashboard 2020 [updated 9/9/2020. Available from: https://gisanddata.maps.arcgis.com/apps/opsdashboard/index.html#/bda7594740fd40299423467b48e9ecf6.

2. Chinazzi M, Davis JT, Ajelli M, Gioannini C, Litvinova M, Merler S, et al. The effect of travel restrictions on the spread of the 2019 novel coronavirus (COVID-19) outbreak. Science. 2020;368(6489):395–400.

3. European Centre for Disease Prevention and Control Situation dashboard – COVID-19 cases in Europe and worldwide 2020 [updated 23/07/2020. Available from: https://qap.ecdc.europa.eu/public/extensions/COVID-19/COVID-19.html.

4. Ridenhour B, Kowalik JM, Shay DK. Unraveling r 0: Considerations for public health applications. American journal of public health. 2018;108(S6):S445–S54.

5. Komenda M, Bulhart V, Karolyi M, Jarkovský J, Mužík J, Májek O, et al. Complex Reporting of the COVID-19 Epidemic in the Czech Republic: Use of an Interactive Web-Based App in Practice. Journal of Medical Internet Research. 2020;22(5):e19367.

6. Holmdahl I, Buckee C. Wrong but useful—what covid-19 epidemiologic models can and cannot tell us. New England Journal of Medicine. 2020.

7. Wu JT, Leung K, Leung GM. Nowcasting and forecasting the potential domestic and international spread of the 2019-nCoV outbreak originating in Wuhan, China: a modelling study. The Lancet. 2020;395(10225):689–97.

8. Kucharski AJ, Russell TW, Diamond C, Liu Y, Edmunds J, Funk S, et al. Early dynamics of transmission and control of COVID-19: a mathematical modelling study. The lancet infectious diseases. 2020.

9. Backer JA, Klinkenberg D, Wallinga J. Incubation period of 2019 novel coronavirus (2019-nCoV) infections among travellers from Wuhan, China, 20–28 January 2020. Eurosurveillance. 2020;25(5):2000062.

10. Nishiura H, Linton NM, Akhmetzhanov AR. Serial interval of novel coronavirus (COVID-19) infections. International journal of infectious diseases. 2020.

11. Hellewell J, Abbott S, Gimma A, Bosse NI, Jarvis CI, Russell TW, et al. Feasibility of controlling COVID-19 outbreaks by isolation of cases and contacts. The Lancet Global Health. 2020.

12. Giordano G, Blanchini F, Bruno R, Colaneri P, Di Filippo A, Di Matteo A, et al. Modelling the COVID-19 epidemic and implementation of population-wide interventions in Italy. Nature Medicine. 2020:1–6.

13. Zhao S, Lin Q, Ran J, Musa SS, Yang G, Wang W, et al. Preliminary estimation of the basic reproduction number of novel coronavirus (2019-nCoV) in China, from 2019 to 2020: A data-driven analysis in the early phase of the outbreak. International journal of infectious diseases. 2020;92:214–7.

14. Zhang S, Diao M, Yu W, Pei L, Lin Z, Chen D. Estimation of the reproductive number of novel coronavirus (COVID-19) and the probable outbreak size on the Diamond Princess cruise ship: A data-driven analysis. International journal of infectious diseases. 2020;93:201–4.

15. Guan W-j, Ni Z-y, Hu Y, Liang W-h, Ou C-q, He J-x, et al. Clinical characteristics of coronavirus disease 2019 in China. New England journal of medicine. 2020;382(18):1708–20.

16. Tindale L, Coombe M, Stockdale JE, Garlock E, Lau WYV, Saraswat M, et al. Transmission interval estimates suggest pre-symptomatic spread of COVID-19. MedRxiv. 2020.

17. Nishiura H, Kobayashi T, Miyama T, Suzuki A, Jung S-m, Hayashi K, et al. Estimation of the asymptomatic ratio of novel coronavirus infections (COVID-19). International journal of infectious diseases. 2020;94:154.

